# Circulating biomarkers of bronchoalveolar injury help predict the need for mechanical ventilation in patients with moderate to severe COVID-19 pneumonia: a prospective cohort study

**DOI:** 10.1101/2025.11.16.25340358

**Authors:** Jérôme Allardet-Servent, Nathalie Hezard, Christel Pissier, Nathalie Bardin, Frédéric Cohen, Aurélie Dehaene, Rettinavelou Soundaravelou, Philippe Halfon, Anderson D. Loundou, Marie-Christine Alessi, Pierre-Emmanuel Morange

## Abstract

**Background:** Severe respiratory failure is a major complication of SARS-CoV-2 infection, and the need for mechanical ventilation (MV) is associated with a worse outcome. Whether some soluble biomarkers of lung injury can help predict MV requirement remains unclear.

**Methods:** This prospective, observational, monocentric cohort study consecutively enrolled patients with laboratory-confirmed COVID-19 pneumonia within 48 h of hospital admission. The serum concentrations of five key bronchoalveolar epithelial and endothelial biomarkers were determined at Day 0, 7 and 14: Krebs von den Lungen-6 (KL-6); soluble receptor for advanced glycation end-products (sRAGE); club cell protein 16 (CC16); angiopoietin-2 (Ang-2); and soluble CD146 (sCD146). The respiratory severity of COVID-19 pneumonia was defined by the maximal level of respiratory support received during hospitalization: oxygen (by mask or nasal prong); high flow oxygen therapy (HFOT); and MV. End-points were the need for MV during hospitalization and the time to liberation from oxygen.

**Results:** Fifty-four COVID-19 patients were enrolled; 23 (43%) required MV, 13 (24%) HFOT, and 18 (33%) oxygen. At inclusion, levels of KL-6, sRAGE, and CC16 were significantly higher in MV compared with non-MV patients (p < 0.05), with sRAGE showing the greatest difference (2.4-fold increase). In multivariate logistic regression, sRAGE (OR per 100 pg/mL increase, 1.028 [95% CI, 1.004–1.054]; p = 0.022) and SpO_2_/F_I_O_2_ (OR, 0.984 [95% CI, 0.970–0.998]; p = 0.008) were identified as independent risk factors for MV. Furthermore, patients with an sRAGE ≥ 5449 pg/mL at inclusion had a lower probability of weaning from oxygen at Day 60 (HR, 0.36 [95% CI, 0.19–0.67]; p = 0.001). From Day 7 to Day 14, CC16 levels increased while sCD146 levels decreased in MV patients.

**Conclusion:** Among five circulating biomarkers of bronchoalveolar injury, sRAGE showed the most favorable kinetic profile, rapidly increasing in MV patients. The early measurement of sRAGE and SpO_2_/F_I_O_2_ upon hospital admission may effectively identify COVID-19 patients at high risk of requiring MV and prolonged oxygen support.

## Introduction

Lung infection caused by the initial variants of severe acute respiratory syndrome coronavirus 2 (SARS-CoV-2), known as coronavirus disease 2019 (COVID-19), was most commonly associated with mild to moderate symptoms, but some patients developed severe hypoxemia and acute respiratory distress syndrome (ARDS) [1]. Predisposing risk factors for severe disease included older age, male sex, obesity, diabetes, hypertension, and chronic organ disease [2–5]. Among patients hospitalized with COVID-19 pneumonia, around 20% required admission to the intensive care unit (ICU) and those requiring mechanical ventilation (MV) had a worse outcome [3,6,7].

SARS-CoV-2 invades the bronchial epithelial cells, type I and type II alveolar pneumocytes, and capillary endothelial cells, ultimately causing cell death [8]. Monitoring circulating biomarkers, released by specific cell subtypes in response to damage, represents a noninvasive opportunity to quantify the extent of lung injury and predict the clinical severity of the disease [9].

The receptor for advanced glycation end-products (RAGE) is a transmembrane pattern recognition receptor constitutively expressed by type I pneumocytes [10]. The soluble form (sRAGE) is derived mainly from cleavage of the extracellular domain, and sRAGE levels indicate alveolar damage in ARDS [11,12]. In COVID-19 patients, sRAGE levels increase in relation to the clinical severity [13–17]. Krebs von den Lungen-6 (KL-6), a specific epitope of the high-molecular-weight glycosylated mucin 1 protein, is secreted by regenerating type II alveolar and bronchial epithelial cells to regulate fibroblast proliferation, migration, and survival [18,19]. KL-6 levels also increase in patients with severe COVID-19 pneumonia [20–27]. Club cell protein 16 (CC16) is a small secretory protein released by non-ciliated bronchial epithelial cells to mitigate the inflammatory response and facilitate epithelial regeneration [28–30]. A delayed increase in CC16 levels has been reported in severe COVID-19 cases [15,31].

Angiopoietin-2 (Ang-2) is an angiogenic factor released by endothelial cells upon activation [32]. COVID-19 patients with a delayed elevation of Ang-2 levels experience worse outcomes [15,33,34]. CD146 is an endothelial cell adhesion molecule involved in vessel integrity and cell migration [35,36]. Cleavage of the extracellular domain of the transmembrane glycoprotein liberates its soluble form (sCD146) [37]. One study reported higher plasma sCD146 levels in COVID-19 patients compared with healthy controls [38].

Although several studies have focused on circulating biomarkers of lung injury during COVID-19, none have compared these three prominent biomarkers of epithelial injury (i.e., sRAGE, KL-6, and CC-16), and the two biomarkers of endothelial injury (Ang-2 and sCD146). The aims of the current study were to describe the kinetics of these biomarkers in relation to disease severity and to determine whether these biomarkers could help predict the need for MV. Our hypothesis was that a combination of biomarkers with other parameters would reliably predict the requirement for MV. Some of the results of this study were presented in an abstract form at the *French Intensive Care Society International Congress* REANIMATION 2022 (FC-250) [39].

## Methods

This prospective, observational, cohort study was conducted at the European Hospital of Marseille, from 26^th^ November 2020 to 08^th^ June 2021, in accordance with the Declaration of Helsinki and the French law on research involving humans. The study protocol was approved by an independent national review board (Comité de Protection des Personnes, Ile de France XI, IDRCB 2020-A00756-33) and registered at ClinicalTrials.gov (NCT04816760). All patients signed written informed consent prior to inclusion. The reporting of this study follows the STROBE recommendations for cohort studies [40].

### Patient selection and management

Adult patients with symptomatic SARS-CoV-2 infection were eligible for inclusion in the study if they fulfilled the following criteria: (i) hospitalized for ≤48 h; (ii) a positive SARS-CoV-2 reverse transcriptase-polymerase chain reaction test; (iii) evidence of COVID-19 pneumonia on a chest computed tomography (CT) scan (bilateral ground-glass opacities and/or consolidations); and (iv) acute onset of respiratory symptoms (≤1 week). The exclusion criteria were: pregnancy; immunosuppressive therapies within the previous 3 months; chronic respiratory failure with home oxygen or non-invasive ventilation; patients with a “do not resuscitate” order or an expected lifespan of <72 h; patients referred from another center with a length of stay >48 h; and patients unable to give informed consent.

Patients with COVID-19 pneumonia were managed according to the World Health Organization (WHO) living guideline, including standard anticoagulation in the absence of thrombosis, corticosteroid therapy (dexamethasone, 6 mg/day for 10 days) if requiring supplemental oxygen, and interleukin-6 receptor blocker therapy (tocilizumab, 8 mg/kg up to a maximum of 800 mg) in severe or critical illness. COVID-19 patients undergoing MV received low tidal volume protective ventilation, prone positioning if PaO_2_/F_I_O_2_ was ≤150 mmHg, and venovenous extracorporeal membrane oxygenation (ECMO) in accordance with guidelines. Patients who developed clinical features of lung fibrosis also received rescue corticosteroid therapy (methylprednisolone, 2 mg/kg/day for 14 days). Twenty healthy individuals who regularly donated blood to the Etablissement Français du Sang served as contemporary controls.

### Samples and laboratory analysis

Blood samples were collected once from controls and at three time points (Day (D) 0, 7, and 14) from COVID-19 patients. The first sample (D0) was collected on the day of inclusion, which occurred within the first 48 h following hospital admission. The concentrations of five biomarkers of bronchoalveolar epithelial and endothelial lung injury were measured in the serum of controls and COVID-19 patients: KL-6; sRAGE; CC16; Ang-2; and sCD146. Whole blood samples were collected in SST™ II Advance BD Vacutainer® tubes, centrifuged for 10 min at 2300 × g, and serum was separated and immediately stored at −80°C for delayed analyses. Commercially available enzyme-linked immunosorbent assay (ELISA) kits from R&D Systems (Quantikine^®^, Minneapolis, MN, USA) were used to quantify sRAGE, CC16, and Ang-2, and from Biocytex (Cy-Quant^®^, Marseille, France) to quantify sCD146. All ELISA measurements were performed with the same lot number on thawed serum samples following recommendations from the manufacturer; the mean values of duplicated assays are reported. The accuracy of ELISA was assessed using replicates of the same control serum positioned at different locations within each plate. KL-6 levels were determined using a chemiluminescent enzyme immunoassay on a Lumipulse^®^ G1200 analyzer (Fujirebio, Tokyo, Japan). Laboratory indices assessed were complete blood count, neutrophil/lymphocyte ratio (NLR), creatinine, C-reactive protein (CRP), ferritin, lactate dehydrogenase (LDH), and D-dimer.

### Lung opacities assessment

Images from chest high resolution computed tomography (HRCT) were processed with *syngo*.via CT Pneumonia Analysis software (Version 1.0.4.2, Siemens Healthineers, Forchheim, Germany). This fully automated AI-based software delineates airspace opacities and provides lobe-wise opacity scores, percentage of opacity (relative to overall lung volume), and percentage of high opacity (relative to overall lung volume). To distinguish between ground glass opacities and consolidations, a Hounsfield Unit (HU) threshold of −200 HU was applied inside the detected airspace opacities. Areas denser than −200 HU were considered consolidations and classified as high opacities.

The opacity score was calculated according to the method reported by Bernheim et al. [41].

Each scan was evaluated for the mean total HU (whole lung parenchyma), opacity score (range: 0–20), percentage of opacity (range: 0–100%), and percentage of high opacity (range: 0–100%). A final quality control by two experienced radiologists, including manual review and editing, was performed to complete the lung segmentations. The performance of the algorithm has been reported elsewhere [42,43].

### Data collection and outcomes

The main demographic characteristics, comorbidities, medical history, length of hospital and ICU stays, types and duration of organ support, and hospital survival were recorded. The Simplified Acute Physiology Score II (SAPS II) was obtained at hospital admission [44], and the Sequential Organ Failure Assessment (SOFA) score was calculated on the days of measurements [45]. Respiratory severity was defined by the type of respiratory support on the World Health Organization clinical progression scale (WHO-CPS) [46] including: oxygen by mask or nasal prong [WHO-CPS grade 5]; high flow oxygen therapy (HFOT) [WHO-CPS grade 6]; and MV [WHO-CPS grade 7–9]. Oxygen-free days were defined as the number of days alive and free from oxygen therapy between inclusion and the time point of interest. End-points were the requirement for MV during the hospital stay and weaning from oxygen at D28 and D60.

### Statistical analysis

The sample size was determined assuming a proportion of patients requiring mechanical ventilation of 30% [47], a margin of error of 10%, and a confidence level of 90% in a population of 1000 subjects. Fifty-four subjects were needed to estimate the true population proportion. Twelve missing data (1.2%), most likely missing at random, for five variables (four for ferritin, two for lactate dehydrogenase, two for D-dimer, two for % of opacity, and two for % of high opacity) were imputed using the multivariate imputation by chained equation (mice) R statistical package (R 4.4.1, R Core Team, 2024) [48].

Categorical data are presented as number and percentage (%), and were compared using the Chi-square test or Fisher’s exact test. Continuous data are presented as median and interquartile range [IQR 25–75%], and were compared using nonparametric tests. Comparisons between controls and COVID-19 subgroups were performed using the Kruskal-Wallis test followed by Conover’s test for *post-hoc* multiple comparisons. Data for COVID-19 patients stratified by MV were compared using the Mann-Whitney U test and Odds ratios (ORs) with 95% confidence intervals [95% CI] were determined by univariate logistic regression. The performance of the biomarkers to predict MV and weaning from oxygen was assessed by the area under curve (AUC) through receiver operating characteristic (ROC) analysis, and the criterion was determined by the Youden index method. Risk factors for MV were determined by multivariate stepwise logistic regression and risk factors for weaning from oxygen were determined using Cox proportional-hazard regression. The time to weaning from oxygen was compared using the log rank test and was represented with Kaplan-Meier curves. The correlations between variables were assessed using Spearman’s coefficient of rank correlation (ρ). Biomarker kinetics were investigated using generalized linear model analyses (GLM) to test the effect of time (D0, 7, and 14), MV (binary), and their interaction. All tests were two-tailed and the significance level was set at 5%.

Statistical analyses were carried out using IBM SPSS Statistics (v29.0.2.0 Armonk, NY, USA) and graphics were created using MedCalc Statistical Software (v23.0.2, Ostend, Belgium) and the ggplot2 R package [49].

## Results

### Study population

Fifty-four patients with COVID-19 pneumonia and 20 healthy controls were included in the study. The main baseline characteristics are reported in **Table 1**. The flow chart of the study population is shown in **S1 Fig** in the Appendix. Eighteen COVID-19 patients (33%) were managed on the ward and received only oxygen therapy during hospitalization. Thirty-six COVID-19 patients (67%) required ICU admission; of these 13 (24%) received HFOT and 23 (43%) required MV. Among the 23 MV patients, 21 (91%) received prone positioning, four (17%) required venovenous ECMO, 18 (78%) received norepinephrine, and five (9%) needed renal replacement therapy.

**Table 1.**
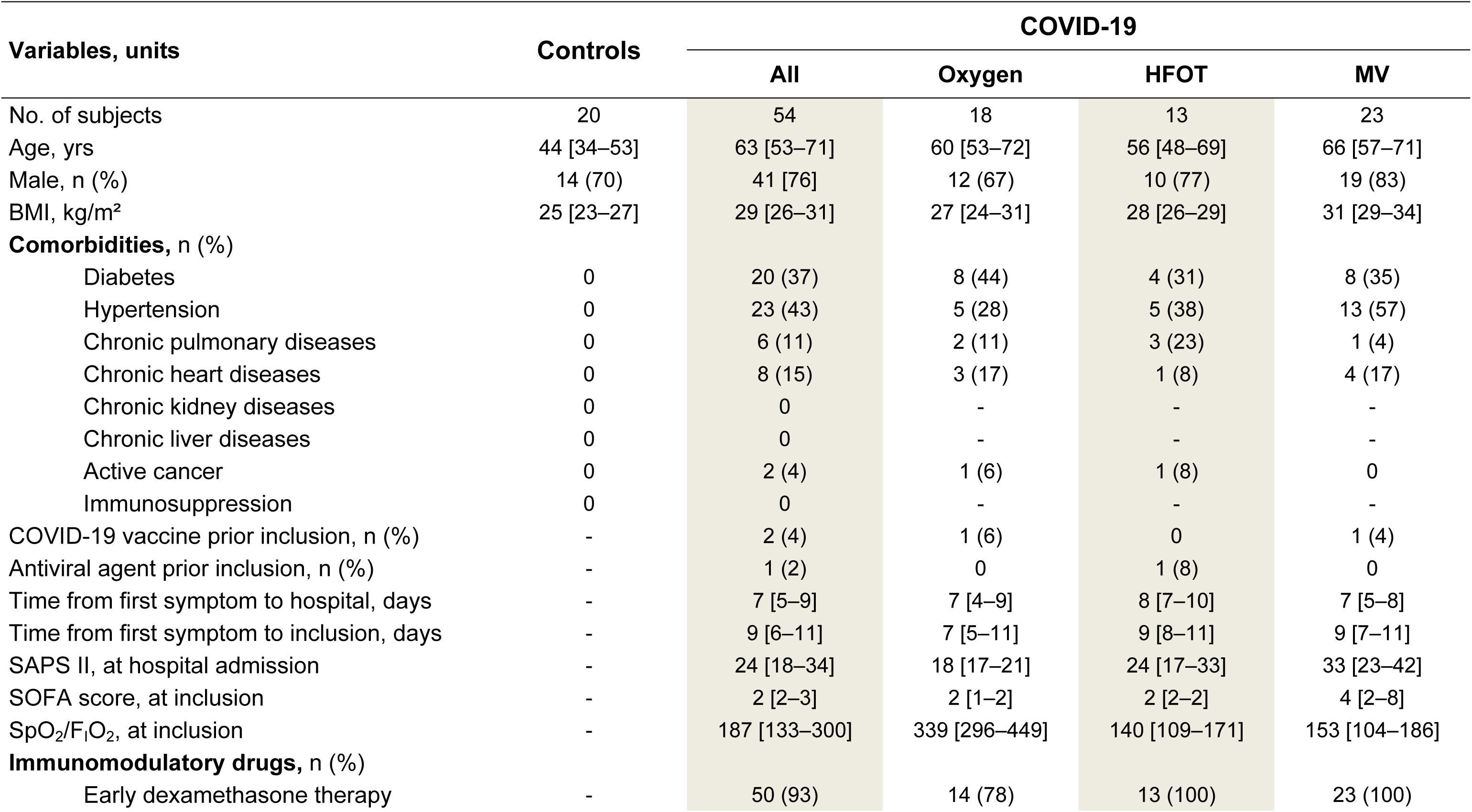
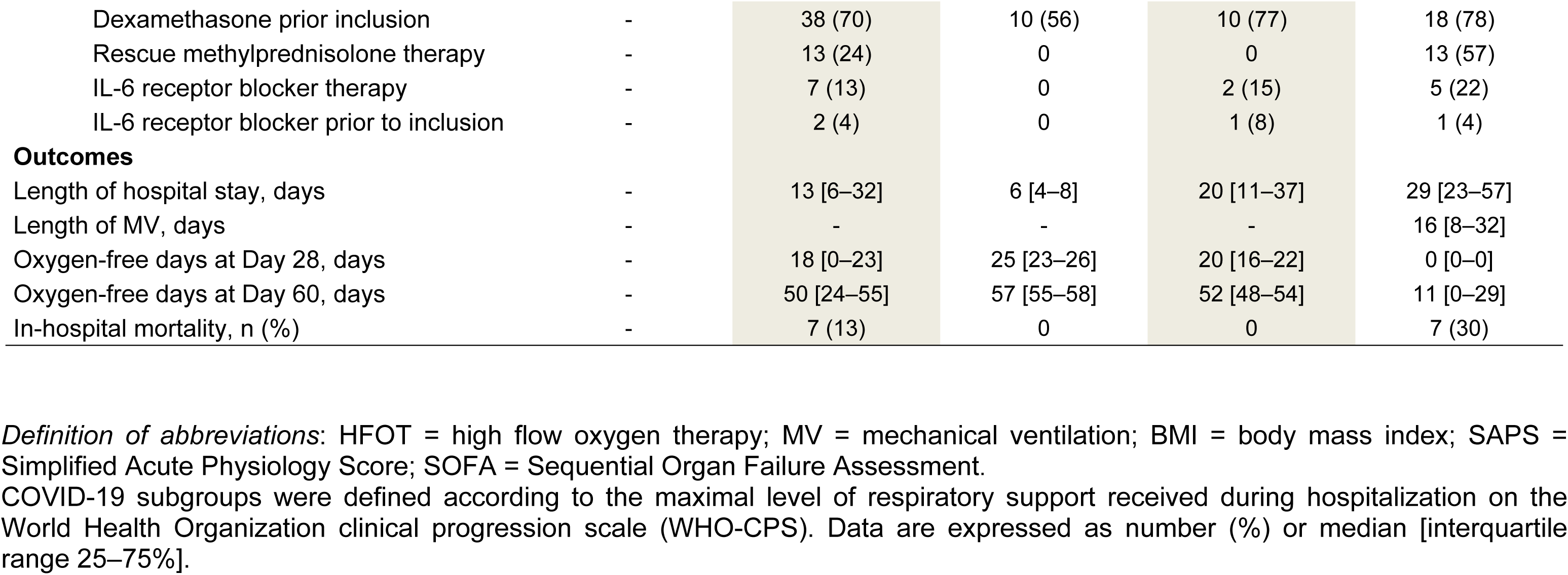
Baseline characteristics of control subjects and COVID-19 patients.

COVID-19 subgroups were defined according to the maximal level of respiratory support received during hospitalization on the World Health Organization clinical progression scale (WHO-CPS). Data are expressed as number (%) or median [interquartile range 25–75%].

### Biomarkers, laboratory, CT, and clinical parameters by respiratory support

The median delay between hospital admission and the first blood sample (D0) was 1 [1;2] day. The serum levels of bronchoalveolar epithelial and endothelial biomarkers are shown in **Fig 1** and values are reported in **S1 Table**. KL-6 levels were higher in the oxygen, HFOT, and MV groups compared with controls, and were higher in the MV group compared with the oxygen group. sRAGE levels were higher in all COVID-19 subgroups compared with controls, and were higher in the MV group compared with the oxygen and HFOT groups. CC16 levels were lower in the oxygen and HFOT groups than in controls and the MV group. Ang-2 levels were higher in the MV group compared with controls. sCD146 levels were lower in all COVID-19 subgroups compared with controls, with no differences observed among COVID-19 subgroups.

**Fig 1.**
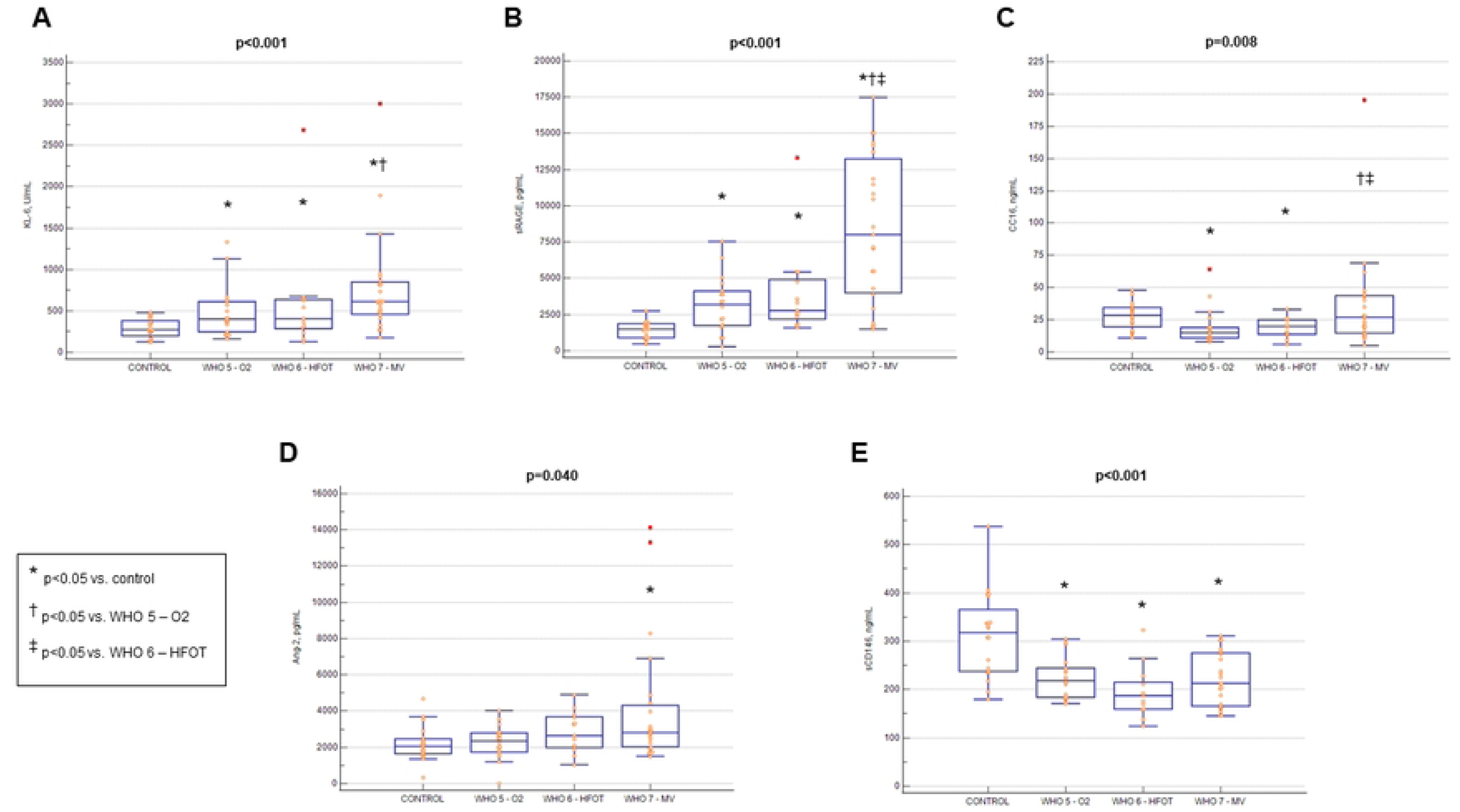
Serum levels of bronchoalveolar epithelial and endothelial biomarkers at inclusion.

Serum samples were obtained within the first 48 h of hospital admission from 54 COVID-19 patients and 20 healthy controls. COVID-19 subgroups were defined according to the maximal level of respiratory support received during hospitalization on the World Health Organization clinical progression scale (WHO-CPS): oxygen by mask or nasal prong (O_2_), high flow oxygen therapy (HFOT), and mechanical ventilation (MV). **(A)** Krebs von den Lungen-6 (KL-6). **(B)** soluble receptor for advanced glycation end-products (sRAGE). **(C)** Club cell protein 16 (CC16). **(D)** Angiopoietin-2 (Ang-2). **(E)** soluble CD146 (sCD146). Statistical analyses were performed using the Kruskal-Wallis test and *post-hoc* multiple comparisons with the Conover test.

The results of laboratory indices and CT-related parameters among COVID-19 subgroups are presented in **S2 Table**. Lung CT scans were performed either prior to (n=52) or after (n=2) inclusion, and the median delay between the CT scan and the first blood sample was -2 [-2;-1] days. The quantity of opacities gradually increased with respiratory severity whilst the mean Hounsfield Unit (HU) of the whole lung parenchyma decreased.

### Biomarkers, laboratory, CT, and clinical parameters by mechanical ventilation

Among the 23 COVID-19 patients requiring MV, the median delay between MV initiation and the first blood sample was 0 [-1;2] day. Seven patients received MV prior to inclusion within a median delay of -1 [-2;-1] day. The main characteristics of the COVID-19 patients stratified by MV are presented in **S3 Table.** MV patients had higher body mass index (BMI), SOFA score, and lower SpO_2_/F_I_O_2_ ratio at inclusion. Laboratory indices (NLR, D-dimer, CRP, ferritin, LDH, and creatinine) and CT-related parameters (% of opacity and % of high opacity) were also higher in MV patients.

Among the bronchoalveolar biomarkers, only KL-6, sRAGE, and CC16 levels were increased in MV patients (**Fig 2**). Four variables (NLR, SOFA score, sRAGE, and % of high opacity) exhibited a ≥2-fold increase. Five variables (CRP, CC16, ferritin, Ang-2, and D-dimer) exhibited a 1.5-to 2-fold increase, and one variable (SpO_2_/F_I_O_2_) decreased by 0.5-fold (**S5 Table**).

**Fig 2.**
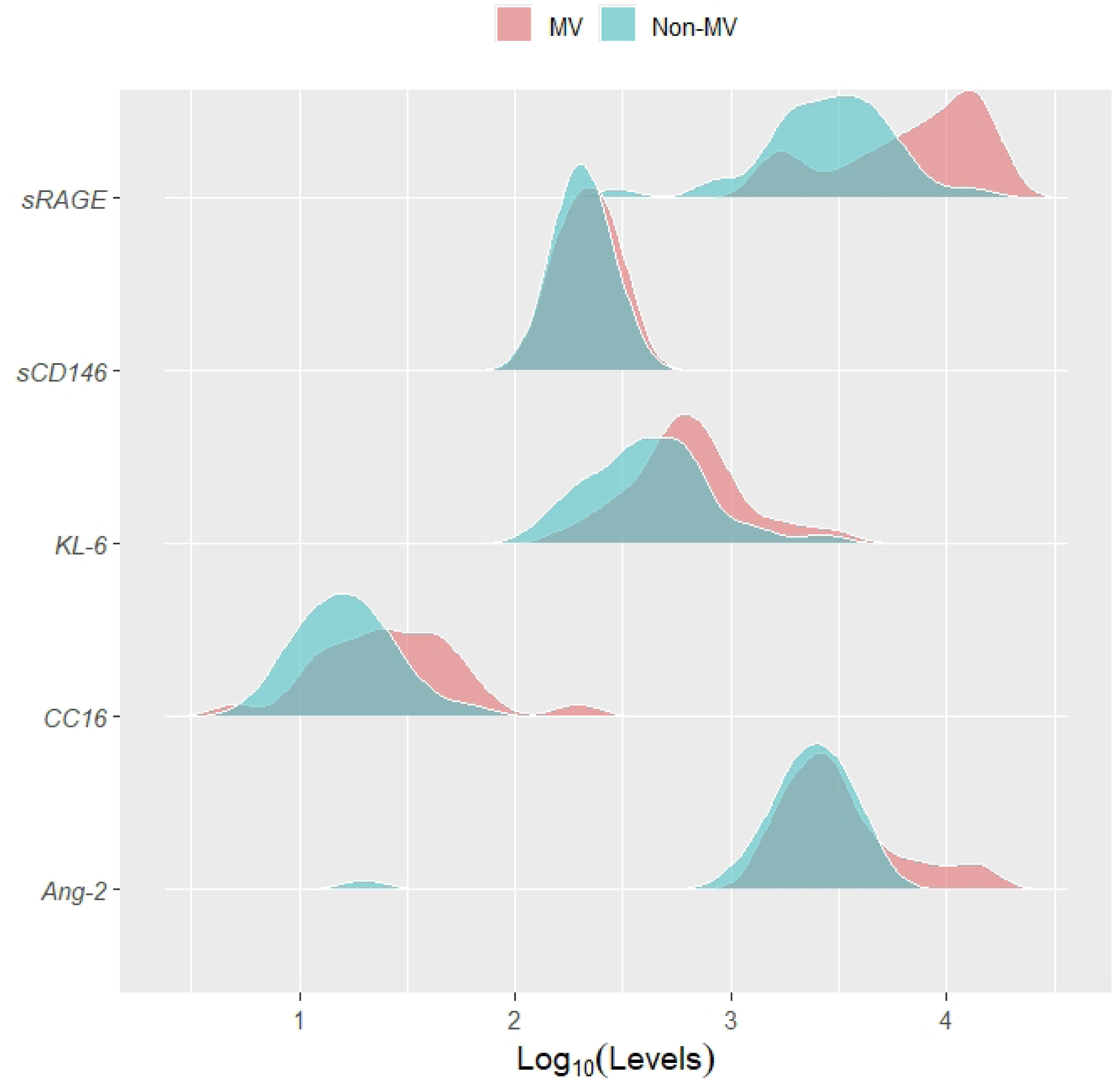
Density distribution of bronchoalveolar biomarkers in COVID-19 patients stratified by mechanical ventilation.

Serum samples were obtained within the first 48 h of hospital admission from 54 COVID-19 patients. The density distribution of five bronchoalveolar epithelial and endothelial biomarkers is represented: Krebs von den Lungen-6 (KL-6), soluble receptor for advanced glycation end-products (sRAGE), club cell protein 16 (CC16), Angiopoietin-2 (Ang-2), and soluble CD146 (sCD146). The distribution of COVID-19 patients who required mechanical ventilation (MV) is colored in red, while those who did not require MV (non-MV) are shown in green. The horizontal axis represents the Log_10_ levels of biomarkers measured in pg/mL for sRAGE and Ang-2, in ng/mL for CC16 and sCD146, and in U/mL for KL-6.

The AUCs to predict the requirement for MV are reported in **S6 Table**. The highest AUC was achieved by the SpO_2_/F_I_O_2_ ratio (0.861 [95% CI, 0.740–0.940]; p<0.001) followed by NLR (0.836 [95% CI, 0.710–0.923]; p<0.001). Among the biomarkers, the highest AUC was achieved by sRAGE (0.786 [95% CI, 0.653–0.886]; p<0.001). In a sensitivity analysis excluding the seven patients receiving MV prior to inclusion, the AUC of sRAGE to predict MV was 0.888 ([95% CI, 0.762–0.961]; p<0.001).

The ORs and 95% CI for MV by univariate and multivariate logistic regression are reported in **Table 2**. After adjusting for confounders, the independent risk factors for MV were sRAGE (OR per 100 pg/mL increase, 1.028 [95% CI, 1.004–1.054]; p=0.022) and SpO_2_/F_I_O_2_ ratio (OR, 0.984 [95% CI, 0.970–0.998]; p=0.008). The model correctly classified 79.6% of cases (AUC, 0.899 [95% CI, 0.786–0.964]; p<0.001).

**Table 2.**
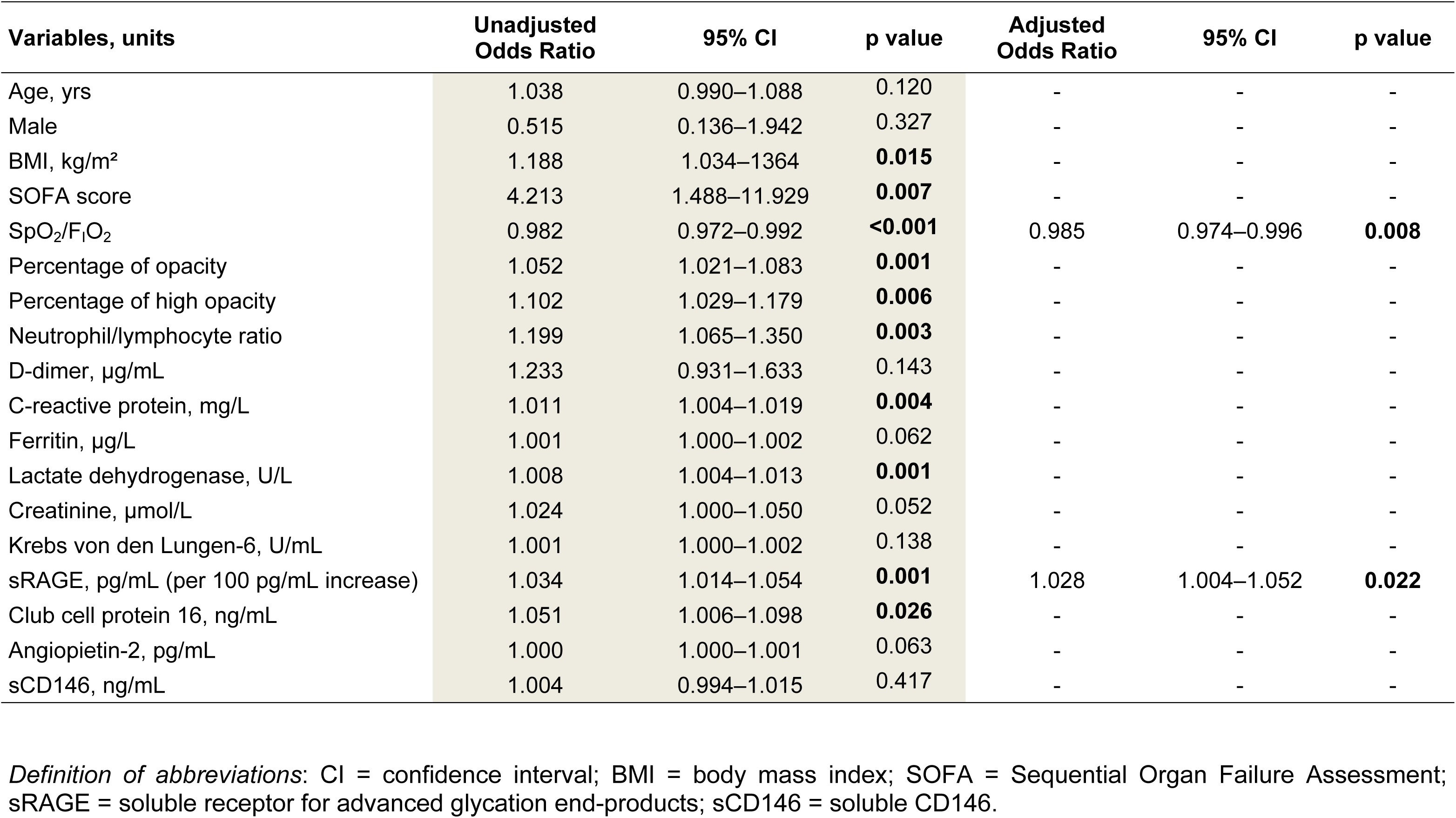
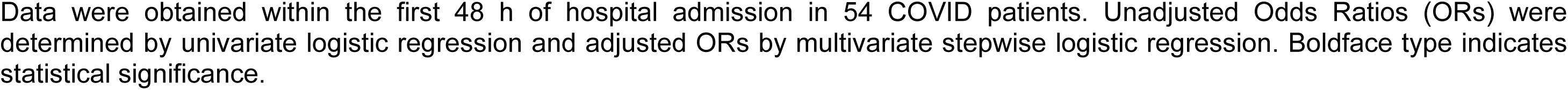
Risk factors for mechanical ventilation in COVID-19 patients.

### Correlation of biomarkers with laboratory, CT, and clinical parameters

The whole set of correlations is summarized through a heat map in **Fig 3**. The biomarkers were statistically associated with several variables. Notably, the percentage of opacity correlated with the levels of sRAGE (ρ=0.475; p<0.001), KL-6 (ρ=0.390; p=0.004), and Ang-2 (ρ=0.307; p=0.027). The percentage of high opacity correlated with Ang-2 (ρ=0.4; p=0.003), KL-6 (ρ=0.358; p=0.009), and sRAGE (ρ=0.356; p=0.009). Negative correlations were observed between the SpO_2_/F_I_O_2_ ratio and the levels of sRAGE (ρ= -0.443; p<0.001), Ang2 (ρ= -0.355; p<0.001), CC16 (ρ= -0.345; p=0.011), and KL-6 (ρ= -0.305; p=0.025). Finally, oxygen-free days at D60 negatively correlated with the levels of CC16 (ρ= -0.448; p<0.001), sRAGE (ρ= -0.413; p=0.002), Ang-2 (ρ= -0.362; p=0.007), and KL-6 (ρ= -0.267; p=0.05).

**Fig 3.**
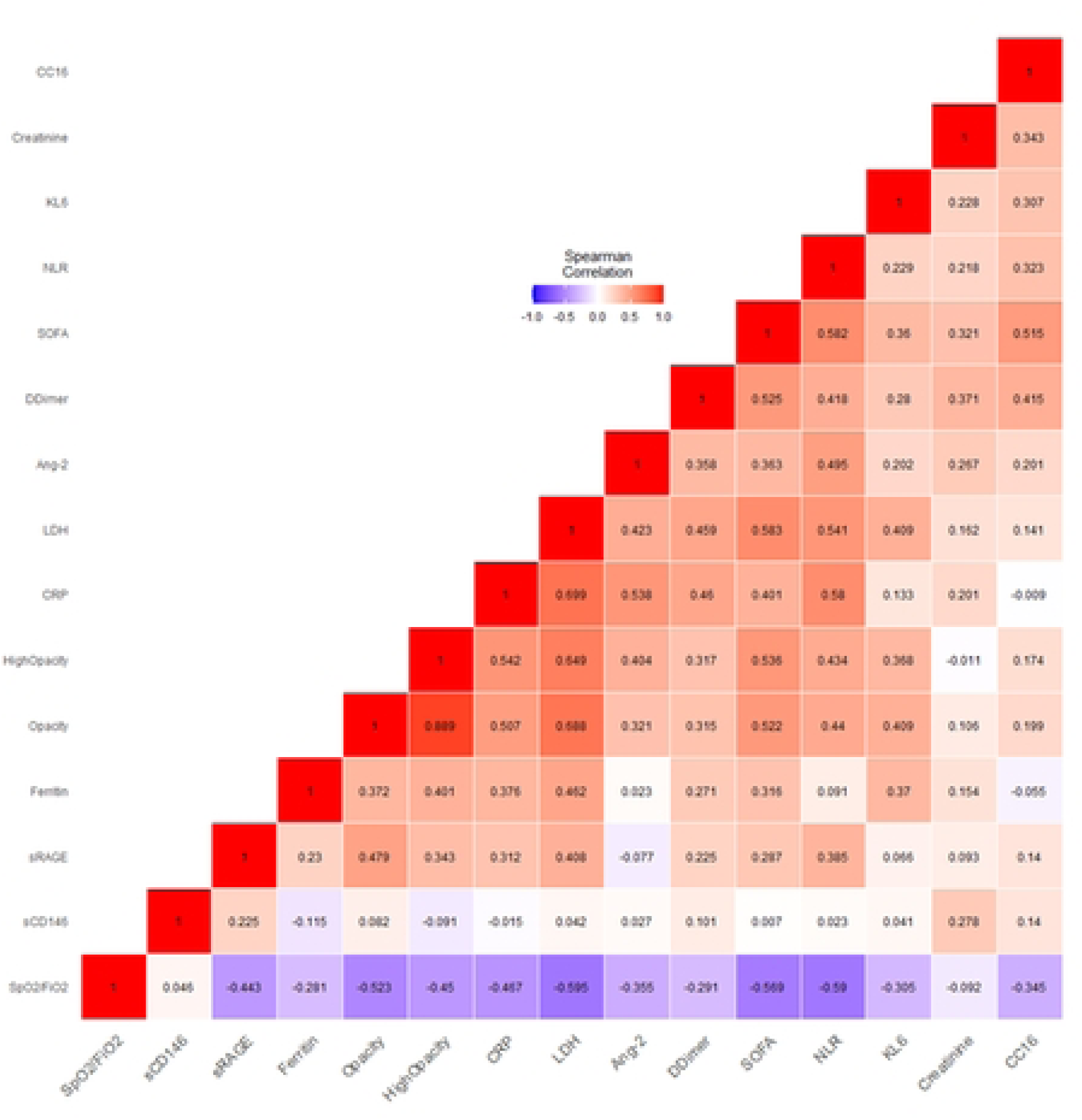
Heat map illustrating biomarker correlations at inclusion in COVID-19 patients.

Measurements were performed within the first 48 h of hospital admission in 54 COVID-19 patients. Correlations were determined between biomarkers of bronchoalveolar epithelial and endothelial injury investigated (Krebs von den Lungen-6 (KL-6), soluble receptor for advanced glycation end-products (sRAGE), Club cell protein 16 (CC16), Angiopoietin-2 (Ang-2), and soluble CD146 (sCD146)), laboratory indices (creatinine, C-reactive protein (CRP), lactate dehydrogenase (LDH), ferritin, D-dimer, and neutrophil/lymphocyte ratio (NLR)), CT-related parameters (percentage of opacity, and percentage of high opacity), respiratory severity index (ratio of peripheral oxygen saturation to inspired oxygen fraction (SpO_2_/F_I_O_2_)), and severity score (Sequential Organ Failure Assessment (SOFA)). Spearman’s rank correlation coefficients are reported.

### Kinetics of biomarkers over 14 days by MV

Complete biomarker kinetics (D0, 7, and 14) were available for 44 COVID-19 patients, among which 22 (50%) received MV (**S7 Table**). The kinetics of KL-6 were time dependent, KL-6 levels being higher at D14 compared with D0 (**Fig 4A**). The kinetics of sRAGE were group and time dependent. sRAGE levels peaked at D0 and were higher in MV patients than in non-MV patients. At D7 and D14, sRAGE levels had decreased compared with D0 and were not different between MV and non-MV patients (**Fig 4B**). The kinetics of CC16 were group and time dependent. CC16 levels increased gradually at each time point in the MV group compared with the non-MV group (**Fig 4C**). The kinetics of Ang-2 were group dependent; Ang-2 levels were higher at D7 in the MV group (**Fig 4D**). The kinetics of sCD146 were group and time dependent; sCD146 levels decreased at D7 and D14 in MV patients (**Fig 4E**).

**Fig 4.**
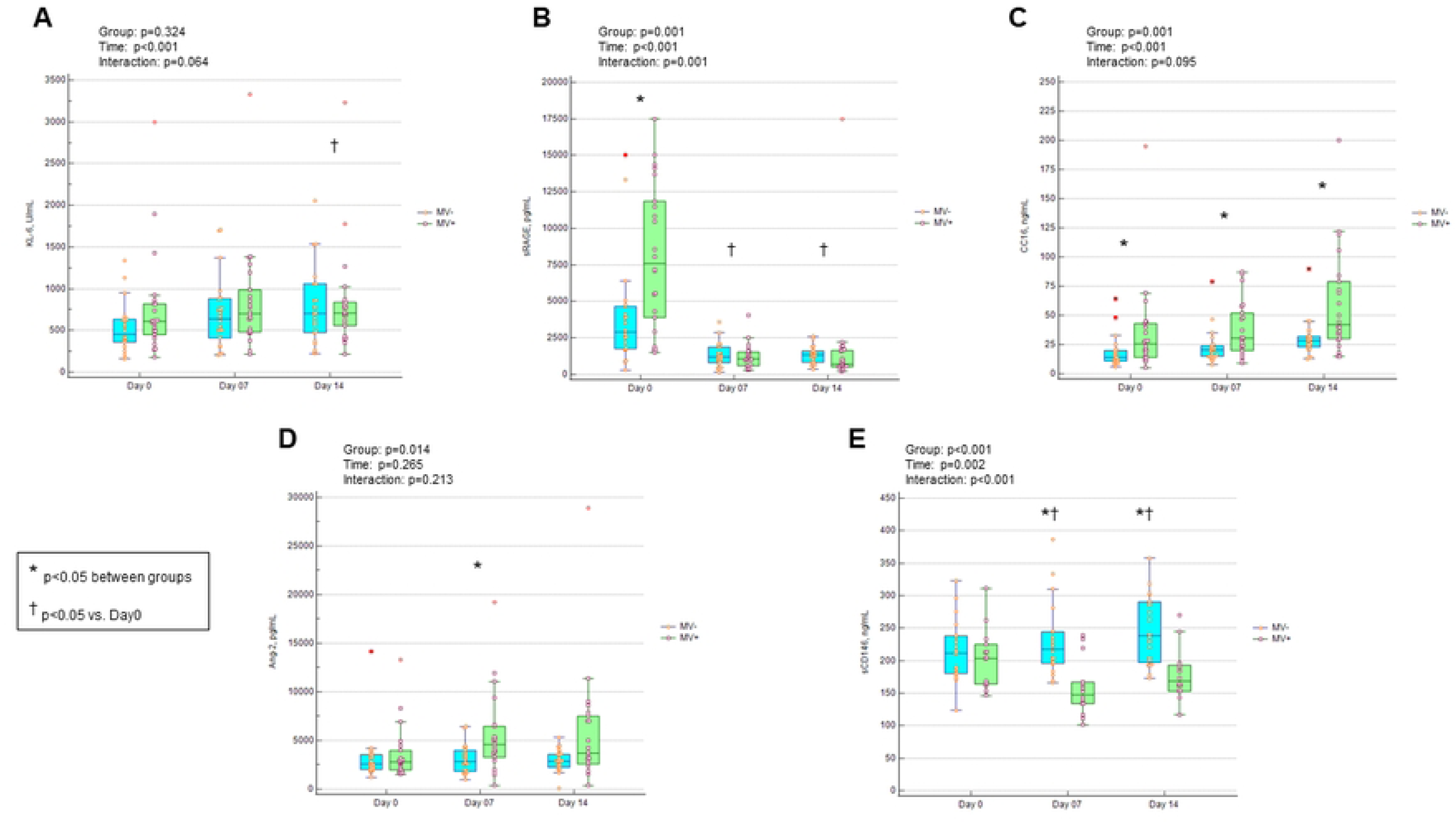
Kinetics of bronchoalveolar epithelial and endothelial biomarkers over 14 days in COVID-19 patients.

Serum samples were obtained from 44 COVID-19 patients at inclusion (Day 0 (D0)) and at D7 and D14 following inclusion. COVID-19 patients were stratified by the need for MV during hospitalization into two groups (MV+ and MV-), both including 22 patients. **(A)** Krebs von den Lungen-6 (KL-6). **(B)** soluble receptor for advanced glycation end-products (sRAGE). **(C)** Club cell protein16 (CC16). **(D)** Angiopoietin-2 (Ang-2). **(E)** soluble CD146 (sCD146). Statistical analyses were performed with generalized linear models (GLM) to test the effect of group (MV), time (day of measurement), and interaction.

### Biomarkers and time to weaning from oxygen

At Day 28, 36 COVID-19 patients (67%) were alive and weaned from oxygen whilst 46 (85%) achieved weaning and seven (13%) had died at D60. At inclusion, sRAGE and CC16 achieved the highest AUCs among the biomarkers to predict weaning from oxygen at D28 (0.796 [95%CI: 0.653–0.886]; p<0.001) and D60 (0.738 [95%CI: 0.600–0.848]; p=0.008), respectively (**S8 and S9 Tables**).

In Cox proportional-hazard regression, four independent factors were associated with the risk of weaning from oxygen at D28: sRAGE (HR per 100 pg/mL increase, 0.989 [95% CI, 0.979–0.999]; p=0.050), SpO_2_/F_I_O_2_ (HR, 1.007 [95% CI, 1.004–1.010]; p<0.001), age (HR, 0.964 [95% CI, 0.938–0.990]; p=0.007), and BMI (HR, 0.881 [95% CI, 0.810–0.957]; p=0.003). COVID-19 patients with an sRAGE level ≥5449 pg/mL at inclusion had a lower probability of weaning from oxygen at D28 (HR, 0.24 [95% CI, 0.12–0.48]; p<0.001, **Fig 5**) and D60 (HR, 0.36 [95% CI, 0.19–0.67]; p=0.001).

**Fig 5.**
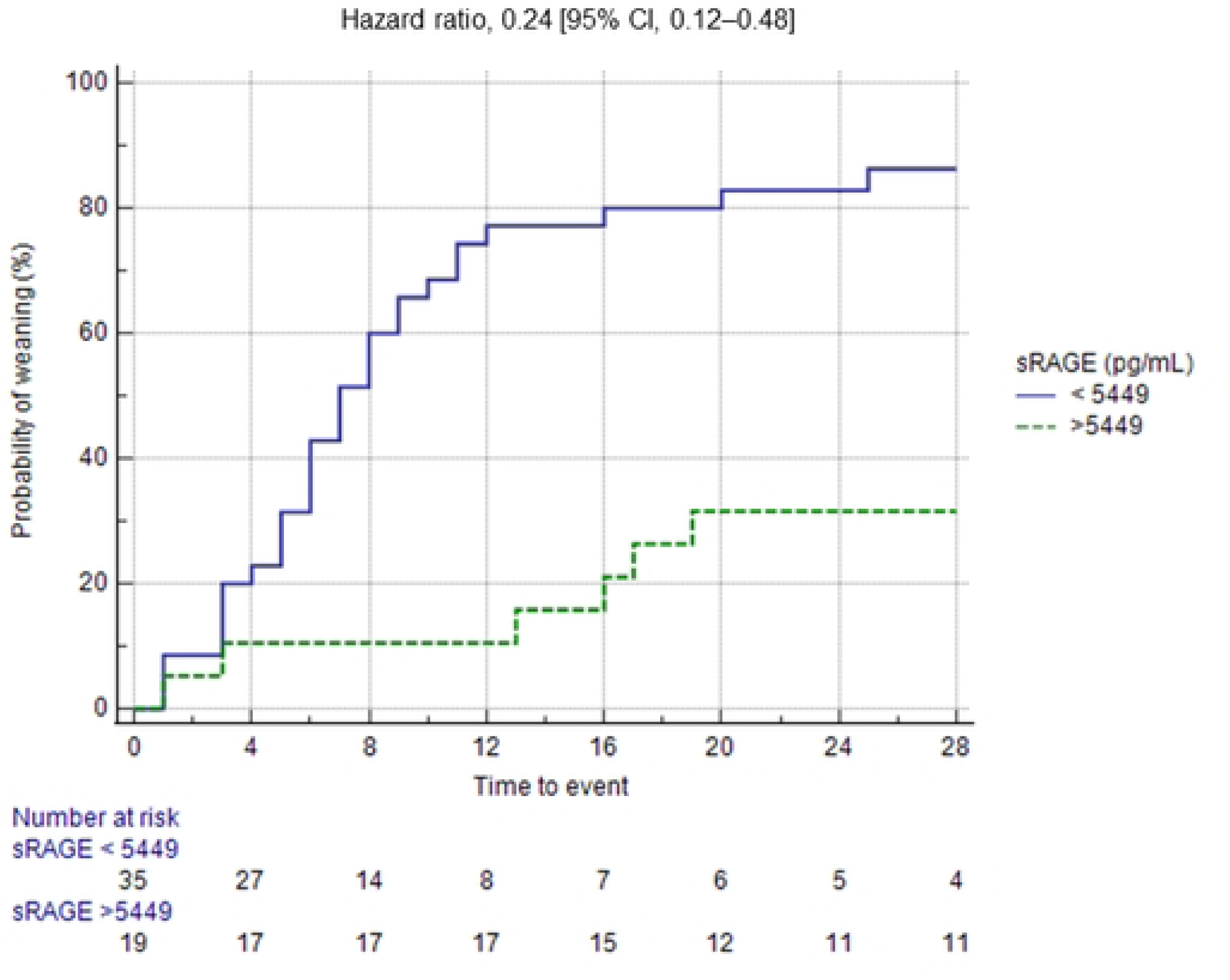
Probability of weaning from oxygen in COVID-19 patients stratified by the sRAGE level at inclusion.

The soluble receptor for advanced glycation end-products (sRAGE) levels were quantified in the serum of 54 COVID-19 patients within the first 48 h of hospital admission. The sRAGE threshold (5449 pg/mL) was determined by the Youden index method. The cumulative probability of weaning from oxygen until Day 28 is reported on the y-axis. Deceased patients (n=7) were censored. Statistical analysis was performed with the Log-rank test to estimate the Hazard ratio (HR) and the 95% confidence interval (CI). Patients with sRAGE levels ≥ 5449 pg/mL had a lower probability of weaning.

## Discussion

This study shows that in hospitalized patients with moderate to severe COVID-19 pneumonia sRAGE had the most favorable kinetic profile among the biomarkers studied with a 2.4-fold increase in MV patients at hospital admission whereas KL-6 was associated with poor discrimination and CC16, Ang-2, and sCD146 had delayed alterations. sRAGE measured within the first 48 h of hospital admission was an independent risk factor for MV and was associated with a lower probability of weaning from oxygen up to D60.

During the course of severe COVID-19 pneumonia, some studies have suggested that circulating biomarkers originating from the alveolar epithelium are released before those from the capillary endothelium [15,50]. Leisman et al. reported a 1.8-fold increase in RAGE levels at admission in the plasma of intubated patients whilst endothelial injury (Ang-2) and Club cell (CC16) biomarkers increased by D3 [15]. The results of our study corroborate and extend these findings. First, we observed a 2.4-fold increase in sRAGE levels at admission in the serum of MV patients that resolved by D7. Second, we confirmed the delayed elevation of Ang-2 and CC16 in MV patients, by D7 in our study. Finally, our results add new findings at D14 showing that CC16 levels continued to increase in MV patients whilst Ang-2 levels decreased. Thus, the kinetic profiles of these biomarkers provide insights into the severity and stage of the disease.

The present study is the first to compare the performance of KL-6 with sRAGE and CC16 in COVID-19 patients. In line with previous reports [20–27], higher KL-6 levels were observed at admission in MV patients, but the discriminating performance of KL-6 was weaker than that of sRAGE. KL-6 levels tended to increase over time without any difference between MV and non-MV patients. Surprisingly, both KL-6 and sRAGE were unaffected by MV status at D7 and D14, in contrast to CC16. Conversely, CC16 levels at hospital admission were poorly discriminant as patients with moderate to severe disease exhibited lower levels than healthy controls. This finding has been reported previously by others [51,52], but the underlying mechanism remains unclear. As a biomarker of bronchoalveolar barrier disruption, the delayed increase in CC16 may reflect either the severity of the initial lung injury and/or the persistence of lung damage through MV [51,53].

This study also provides new insights into sCD146, a biomarker of endothelial activation. A decrease in sCD146 levels was observed at hospital admission in the serum of COVID-19 patients compared with healthy controls, irrespective of disease severity. Furthermore, the kinetics of sCD146 over 14 days depended on MV status; sCD146 levels being much lower in MV patients. These results contrast with those reported by Syed et al. in COVID-19 patients [38]; however, differences in the nature of the samples (plasma vs. serum) and the ELISA assays may have contributed to such discrepancies. Nevertheless, decreased sCD146 levels have already been reported during the acute phase of other inflammatory diseases [54,55]. Thus, the endothelial biomarkers Ang-2 and sCD146 exhibited opposite kinetics over time. Although the levels of these biomarkers were already altered at admission in the most severe patients, endothelial damage worsened at a later stage in MV patients.

Determining the strength of the relation between a biomarker and the extent of lung injury yields valuable information to compare biomarkers, but such data are scarce in patients with COVID-19 pneumonia [56]. In this study, fully automated AI-based software was used to quantify lung opacities. Both the opacity level (semi-quantitative assessment) and the percentage of opacity and high opacity (quantitative assessment) were found to increase gradually with the level of respiratory support. Meanwhile, the mean HU of the whole lung parenchyma decreased gradually, indicating a loss of aeration. LDH achieved the highest level of correlation with both the percentage of opacity and the severity of hypoxemia. With a slightly lower strength, sRAGE was the biomarker of lung injury that exhibited the highest level of correlation with these two parameters.

In hospitalized COVID-19 patients, the risk factors for MV include older age, comorbidities, severe hypoxemia, higher SOFA score, and increased levels of CRP, LDH, D-dimer, creatinine, and NLR [57–59]. However, few studies have included the biomarkers of lung injury. In the present study, sRAGE and the SpO_2_/F_I_O_2_ ratio at inclusion were identified as two independent risk factors for MV after adjustment for confounders. These results corroborate the findings of Lim et al. who identified sRAGE and the SOFA score as risk factors for MV [14]. Overall, sRAGE not only achieved the most favorable kinetic profile with an early increase following hospital admission, but was also an independent risk factor for the prediction of MV. Finally, sRAGE provided additional information on the duration of oxygen supplementation, and patients with increased sRAGE levels at admission had a lower probability of weaning at D60. Further research is needed to determine whether the lung function of these patients remains altered long-term.

This study has some limitations. First, most COVID-19 patients (93%) received corticosteroids during their hospital stay, even prior to inclusion in 69% of cases. Corticosteroids are known to decrease the level of inflammatory markers in blood [60], but the effect on biomarkers of lung injury remains undetermined. Nonetheless, the proportion of patients receiving corticosteroids prior to inclusion was not different between the MV and non-MV groups. Therefore, it is unlikely that corticosteroids contributed to the differences observed in this study. Second, MV was initiated prior the first blood sample in seven of the 23 patients requiring MV, and this could have contributed to the increase in biomarker levels. However, the results were unaffected by excluding these seven patients in the sensitivity analysis. Third, the study was designed as a monocentric determination cohort including a limited number of patients. Therefore, our findings need to be validated in a larger multicentric cohort study. Finally, our results should be restricted to the patients infected by the first variants of the SARS-CoV-2 and treated with dexamethasone.

## Conclusion

In this prospective cohort study investigating the performance of five circulating biomarkers of lung injury in patients hospitalized with moderate to severe COVID-19 pneumonia, a 2.4-fold increase in sRAGE levels was observed in MV patients at hospital admission which resolved by D7. sRAGE and SpO_2_/F_I_O_2_ ratio at inclusion were identified as independent risk factors for MV. Increased sRAGE levels at admission were also associated with a lower probability of weaning from oxygen up to D60. In contrast, KL-6 levels poorly discriminated the need for MV, while Ang-2 and sCD146 shown delayed and opposite alterations in MV patients. CC16 levels increased markedly in MV patients over 14 days, but were close to healthy controls at admission. These findings encourage the measurement of sRAGE at hospital admission to identify patients at risk of MV and prolonged oxygen supplementation.

## Data Availability

https://doi.org/10.5281/zenodo.17550637

## Acknowledgements

We are grateful to Sara Amrani and Adrien Estienne for their invaluable help with the management of blood samples, and to Nadine Para for ELISA measurements. We thank NewMed Publishing Ltd. for English language editing.

## Supporting information

**S1 Fig. Flow chart of the study population.**

Definition of abbreviations: ICU = intensive care unit; O2 = standard oxygen therapy (nasal prong or face mask); HFOT = high flow oxygen therapy; MV = mechanical ventilation.

**S2 Fig. Volcano plot showing fold changes in baseline parameters of COVID-19 patients requiring mechanical ventilation.**

Measurements in COVID-19 patients were performed within the first 48 h of hospital admission. COVID-19 patients were stratified by the need for mechanical ventilation (MV) during hospitalization into two groups. Fold changes (FC) are represented on a log2 scale in the x-axis. False Discovery Rate (FDR) p values are represented on a log10 scale in the y-axis. Labelled variables are those with fold changes <0.5 or >1.5 and FDR p values ≤0.05. SpO_2_/F_I_O_2_ = peripheral oxygen saturation to inspired oxygen fraction ratio; SOFA = Sequential Organ Failure Assessment; NLR = neutrophil/lymphocyte ratio; sRAGE = soluble receptor of advanced glycation end-products; CC16 = Club cell protein 16.

**S1 Table. Serum levels of bronchoalveolar epithelial and endothelial biomarkers at inclusion in healthy controls and COVID-19 patients according to the type of respiratory support.**

Definition of abbreviations: HFOT = high flow oxygen therapy; MV = mechanical ventilation; sRAGE = soluble receptor of advanced glycation end-products; sCD146 = soluble CD146.

Data are presented as median [interquartile range: 25–75%]. Measurements in COVID-19 patients were performed within the first 48 h of hospital admission. Statistical analyses were performed with the Kruskal-Wallis test and post-hoc multiple comparisons with the Conover test. Boldface type indicates statistical significance.

* p<0.05 vs. controls; † p<0.05 vs. oxygen group; ‡ p<0.05 vs. HFOT group.

**S2 Table. Baseline laboratory indices and CT-related parameters in COVID-19 patients according to the type of respiratory support.**

Definition of abbreviations: HFOT = high flow oxygen therapy; MV = mechanical ventilation; HU = Hounsfield unit.

COVID-19 subgroups were defined according to the maximal level of respiratory support received during hospitalization on the World Health Organization clinical progression scale (WHO-CPS). Data are presented as median [interquartile range: 25–75%]. Statistical analyses were performed with the Kruskal-Wallis test and post-hoc multiple comparisons with the Conover test for continuous variables. Boldface type indicates statistical significance.

* p<0.05 vs. oxygen group; † p<0.05 vs. HFOT group.

**S3 Table. Baseline characteristics of COVID-19 patients stratified by the need for mechanical ventilation during hospitalization.**

Definition of abbreviations: BMI = body mass index; SOFA = Sequential Organ Failure Assessment; sRAGE = soluble receptor for advanced glycation end-products; sCD146 = soluble CD146.

Data are expressed as number (percentage) or median [interquartile range: 25–75%]. Statistical analyses were performed with the Mann-Whitney U test for continuous variables and the Chi-square test or Fisher’s exact test for categorical variables. Boldface type indicates statistical significance.

**S4 Table. Receiver operating characteristics analyses of biomarkers, laboratory, CT-related, and clinical parameters at inclusion to predict the need for mechanical ventilation during hospitalization.**

Definition of abbreviations: AUC = area under the curve; CI = confidence interval; Se = sensitivity; Sp = specificity; KL-6 = Krebs von den Lungen-6; sRAGE = soluble receptor of advanced glycation end-products; CC16 = Club cell protein 16; Ang-2 = Angiopoietin-2; sCD146 = soluble CD146; LDH = lactate dehydrogenase; BMI = body mass index; SOFA = Sequential Organ Failure Assessment.

Measurements were performed within the first 48 h of hospital admission in 54 COVID-19 patients. The criterion was determined by the Youden index method. Boldface type indicates statistical significance.

**S5 Table. Serum level kinetics over 14 days of bronchoalveolar epithelial and endothelial biomarkers in COVID-19 patients stratified by the need for mechanical ventilation during hospitalization.**

Definition of abbreviations: MV = mechanical ventilation; KL-6 = Krebs von den Lungen-6; sRAGE = soluble receptor of advanced glycation end-products; CC16 = Club cell protein 16; Ang-2 = Angiopoietin-2; sCD146 = soluble CD146.

Data are presented as median [interquartile range: 25–75%]. Measurements were performed in 44 COVID-19 patients at inclusion (Day 0) and at Day 7 and Day 14 following inclusion. COVID-19 patients were stratified by the need for MV during hospitalization into MV (N=22) and Non-MV (N=22) groups. Statistical analyses were performed with the Kruskal-Wallis test and post-hoc multiple comparisons with the Conover test. Boldface type indicates statistical significance.

* p<0.05 vs. Day 0; † p<0.05 vs. Day 7.

**S6 Table. Receiver operating characteristics analyses of biomarkers, laboratory, CT-related, and clinical parameters at inclusion to predict weaning from oxygen at Day 28.**

Definition of abbreviations: AUC = area under the curve; CI = confidence interval; Se = sensitivity; Sp = specificity; KL-6 = Krebs von den Lungen-6; sRAGE = soluble receptor of advanced glycation end-products; CC16 = Club cell protein 16; Ang-2 = Angiopoietin-2; sCD146 = soluble CD146; LDH = lactate dehydrogenase; BMI = body mass index; SOFA = Sequential Organ Failure Assessment.

Measurements were performed within the first 48 h of hospital admission in 54 COVID-19 patients. The criterion was determined by the Youden index method. Boldface type indicates statistical significance.

**S7 Table. Receiver operating characteristics analyses of biomarkers, laboratory, CT-related, and clinical parameters at inclusion to predict weaning from oxygen at Day 60.**

Definition of abbreviations: AUC = area under the curve; CI = confidence interval; Se = sensitivity; Sp = specificity; KL-6 = Krebs von den Lungen-6; sRAGE = soluble receptor of advanced glycation end-products; CC16 = Club cell protein 16; Ang-2 = Angiopoietin-2; sCD146 = soluble CD146; LDH = lactate dehydrogenase; BMI = body mass index; SOFA = Sequential Organ Failure Assessment.

Measurements were performed within the first 48 h of hospital admission in 54 COVID-19 patients. The criterion was determined by the Youden index method. Boldface type indicates statistical significance.

